# An interactive dashboard to track themes, development maturity, and global equity in clinical artificial intelligence research

**DOI:** 10.1101/2021.11.23.21266758

**Authors:** J Zhang, S Whebell, J Gallifant, S Budhdeo, H Mattie, P Lertvittayakumjorn, M P Arias Lopez, B J Tiangco, J W Gichoya, H Ashrafian, L A Celi, J T Teo

## Abstract

The global clinical artificial intelligence (AI) research landscape is constantly evolving, with heterogeneity across specialties, disease areas, geographical representation, and development maturity. Continual assessment of this landscape is important for monitoring progress. Taking advantage of developments in natural language processing (NLP), we produce an end-to-end NLP pipeline to automate classification and characterization of all original clinical AI research on MEDLINE, outputting real-time results to a public, interactive dashboard (https://aiforhealth.app/).

## Introduction

Interest in the application of artificial intelligence (AI) to human health problems continues to grow, but widespread translation of academic research into deployable AI devices has proven more elusive. There is increasing recognition of limitations in how clinical AI research is conducted^1,2^, from characteristics of data:^3^ to methods for model development4, heterogeneity between clinical specialties in translation to devices^5^, and inadequate inclusion of diverse and global populations^6,7^. Continual quantification of these features can enable identification of shortcomings in a heterogenous landscape while allowing progress to be monitored over time. However, the sheer quantity of published research (more than 150,000 papers on MEDLINE under broad AI-related terms - see *Methods)* makes this a significant challenge. Literature reviews can only map a portion of the research landscape at a single time-point and are laborious to conduct and reproduce. Literature database searches cannot directly identify original research in model development, or pinpoint research that represents advanced stages of model validation.

In response to tremendous growth in AI research publication, we created an end-to-end natural language processing (NLP) pipeline that automates on-going identification, classification, and characterization of original AI research abstracts extracted from the MEDLINE database *(Figure 1)*. Results are output to an interactive dashboard (https://aiforhealth.app), creating a live view of global AI development, browsable by development maturity, medical specialty, data type, algorithm, research location, publication date, or any combination of attributes. Daily updated datasets are made available to download, providing a user-friendly aid to literature reviewers, or for reproducible assessment of research progress.

**Figure 1.**
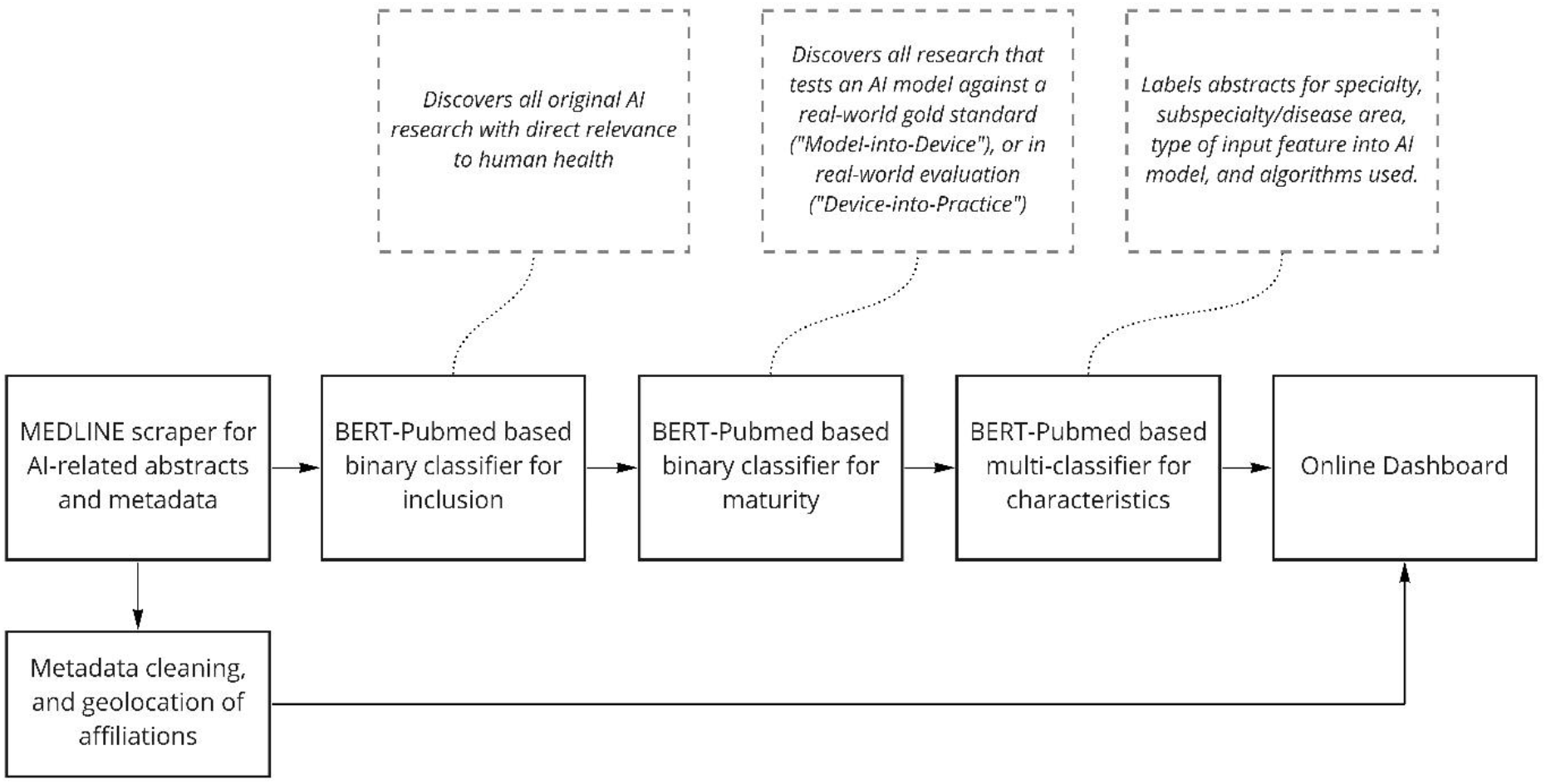
Stages of end-to-end natural language processing pipeline for classifying and characterising all original clinical AI research as indexed on MEDLINE

## Methods

Our aims for the dashboard and pipeline were four-fold. Firstly, to identify original research in clinical AI model development; Second, to identify research at ‘mature’ development stages, describing either evaluation of an AI algorithm vs a reference standard, or prospective real-world *(Figure 2);* Third, to track global distribution and equity of AI research on a per-author basis; Fourth, to characterise, in detail, the main disease areas, clinical specialties, algorithms, and data types in AI research. All development was performed using Python 3.8 and Tensorflow^8^. All manual labelling was performed by JZ, JG and SW.

**Figure 2.**
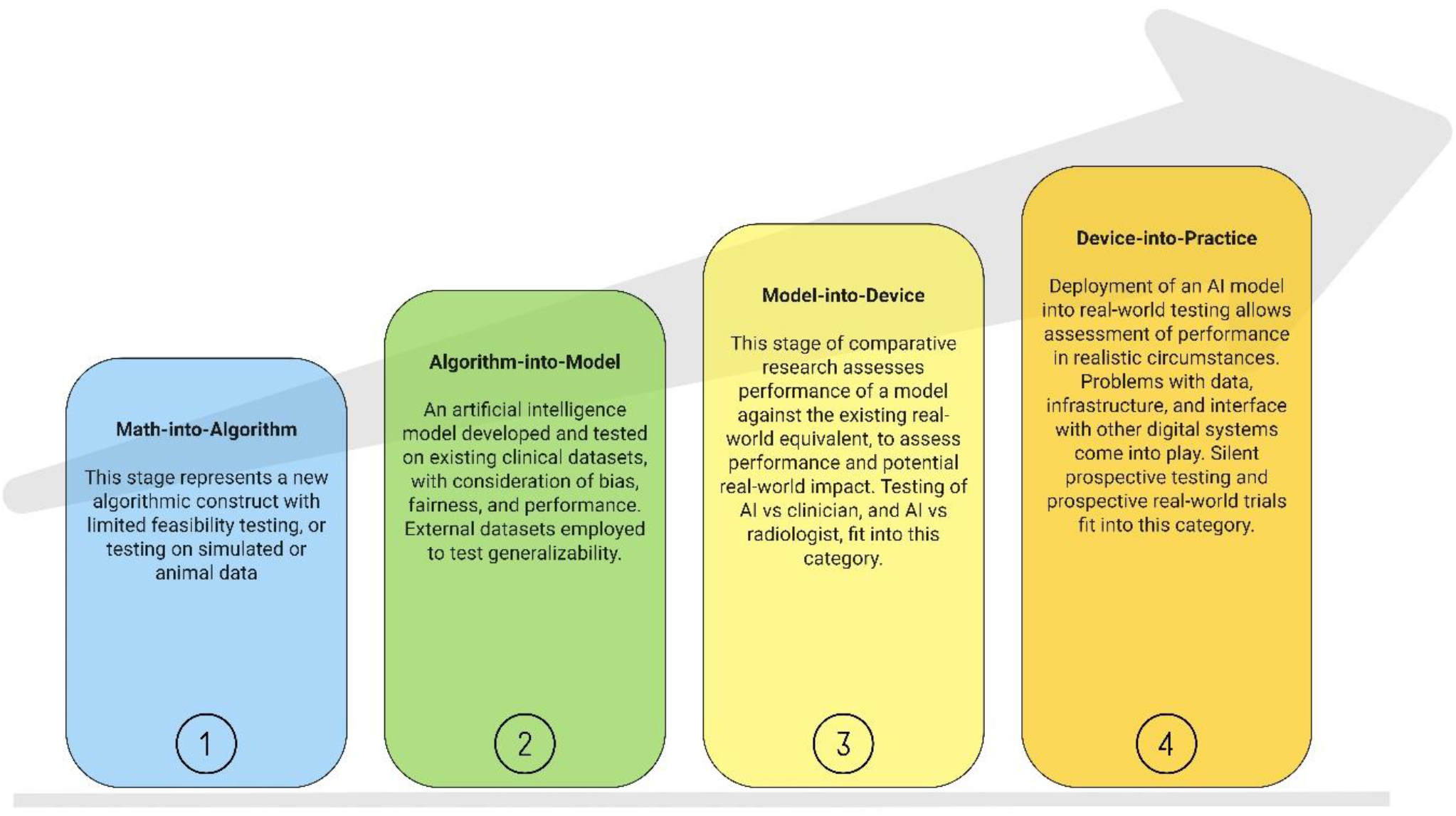
maturity framework used by pipeline

### Publication search and metadata collation

A web-scraping application was produced using Entrez API^9^ to obtain titles, abstracts, metadata, and Medical Subject Heading (MeSH) terms from MEDLINE, for publications under broad AI search terms. From metadata we extracted publication date, journal, authors, affiliations, and derived geographical location of affiliations (see Supplementary Materials).

We used the following search terms for scraping publications from MEDLINE, based on search terms employed in previous systematic reviews of artificial intelligence: “““(((((((([“artificial intelligence”) OR (“deep learning”)) OR (“machine learning”)) OR (“neural net”)) OR (“transfer learning”)) OR (“supervised learning”)) OR (unsupervised learning))) OR (artificial intelligence[MeSH Terms])”““

### Author affiliation geo-location

The location embedded in MEDLINE metadata refers to the journal country, rather than the country where research was conducted. We extracted geographic location of author affiliations using Python 3.8 geocoding libraries, including geopy (v2.2.0) for location, and folium (v0.12.1) for visualization.

### BERT-PubMed

We employed a transfer learning approach, using state-of-the-art Bi-directional Encoder Representations from Transformers (BERT)^10^ NLP models with substantial pre-training on medical corpuses and academic abstracts (BERT-PubMed)^11^.BERT, originally created by Google AI, enables bidirectional text representation to develop a deeper sense of language context. It is resilient to imbalanced datasets without need for additional methods for data augmentation^12^. Classifier models were fine-tuned on manually labelled abstracts indexed on MEDLINE between 1998 and 2020.

Each classifier was trained using a 512-sequence length, increasing accuracy at the expensive of training cost. We used an AdamW optimizer. Training was performed using Tensorflow and Keras libraries on a local machine with an Nvidia GTX graphics card. Optimal epochs ranged from 3, for inclusion and maturity classifiers, to 5 for the multi-classifier. Training data and code hosted at: https://github.com/whizzlah/health_ai_training.

### Inclusion classifier

In determining labelling criteria, we aimed to include all research papers that develop AI models for human healthcare, focusing on models that provide predictive, diagnostic, or quantitatively informative outputs that inform decision-making. We designed a guide question: “Does the proposed model output have a direct, actionable effect on patient care, by providing information to a healthcare provider, patient, or automated system?”. This excludes AI models for pre-processing images or data, or workflow assistance^13^. We aimed to exclude publications using non-human participants, reviews, and informal publication types.

BERT-PubMed was fine-tuned with a binary classification layer. Training was initiated on 4000 abstracts from 1998 to 2020 manually labelled for inclusion. Negligible numbers of positive samples were found prior to 1998. The training set was augmented actively by manually labelling additional abstracts classified with high uncertainty, until model performance reached satisfactory metrics. Evaluation was conducted on a test set (n=1034), and prospectively on abstracts from publications produced after pipeline completion (n=lO00, after September 2021). Classifier sensitivity against manual review was tested using a curated list of publications (n=446) from a recent systematic review of deep learning^14^ (*Table 1)*.

**Table 1.** evaluation metrics for BERT-PubMed inclusion classifier

|  |  |
| --- | --- |
| **Test set (n=1034)** |  |
| Precision | 0.94 |
| Recall | 0.96 |
| Specificity | 0.98 |
| Accuracy | 0.98 |
| F1-Score | 0.95 |
| Matthew’s correlation coefficient | 0.94 |
| **Prospective 2021 abstracts (n=1000)** |  |
| Precision | 0.95 |
| Recall | 0.96 |
| Specificity | 0.98 |
| Accuracy | 0.98 |
| F1-Score | 0.96 |
| Matthew’s correlation coefficient | 0.94 |
| **Curated papers from systematic reviews of AI (n=446)** |  |
| Publications correctly labelled by model | 438 |
| Sensitivity | 0.98 |

### Maturity classifier

Previous publications examine AI ‘maturity’ in two separate but parallel contexts^15,16^. The first describes important methodological, data, and reporting characteristics within model training and testing that contribute to risk of bias and calibration. The second considers a high-level overview of stages of technological development, with previous research adapting National Aeronautics and Space Administration (NASA) technology readiness. However, these were designed for prototyping engineering devices and do not necessarily translate to development of heterogenous clinical AI models.

We consider technological development maturity using a novel framework specific to clinical AI, describing four stages prior to active real-world implementation (Figure 2). In summary, a “Math-into-Algorithm” stage represents development of novel algorithmic techniques, an “Algorithm-into-Model” stage represents the testing of model performance on datasets with ground truth labels, a “Model-into-Device” stage describes model testing against a non-AI, existing, gold standard (analogous to a comparative clinical study), and “Device-into-Practice” describes deployment for validation in a prospective, real-world environment. This framework has been employed in a recent review of AI in mechanical ventilation^17^.

We fine-tuned a second BERT-PubMed classifier to identify abstracts fulfilling at least a “Model-into-Device” stage of model development, initiated on 2500 manually labelled abstracts from 1998 to 2020 with active augmentation. The maturity classifier was evaluated on a test set (n=784), prospectively on abstracts from 2021 (n=2494, after September 2021), and output compared to curated publications from a systematic review of AI vs clinicians^18^ (n=83) (*Table 2)*.

**Table 2.** evaluation metrics for BERT-PubMed maturity classifier

|  |  |
| --- | --- |
| **Test set (n=784)** |  |
| Precision | 0.91 |
| Recall | 0.96 |
| Specificity | 0.99 |
| Accuracy | 0.99 |
| F1-Score | 0.93 |
| Matthew’s correlation coefficient | 0.93 |
| **Prospective 2021 abstracts (n=2494)** |  |
| Precision | 0.94 |
| Recall | 0.89 |
| Specificity | 0.99 |
| Accuracy | 0.99 |
| F1-Score | 0.91 |
| Matthew’s correlation coefficient | 0.90 |
| **Systematic review of AI vs clinician (n=83)** |  |
| Publications correctly labelled by model | 81 |
| Sensitivity | 0.97 |

### Study characteristic labelling and classification

We used a Named-Entity-Recognition (NER) NLP model (SparkNLP based on work by Chiu/Nichols) combined with a dictionary-based text recognition layer to discover and classify major entities expressed in abstracts, including clinical specialty, subspecialty or disease, type of algorithm used, and type of data input into models. For ease of interpretation, we did not consider Radiology as a ‘clinical specialty’, instead considering use of radiomics as an input feature. This avoids potential inconsistencies where, for example, an AI device for stroke diagnosis could either be classified under Radiology or Neurology. For the purposes of online deployment, manually validated NER labels were used to train a BERT-PubMed multi-classifier to label abstracts for major specialties, subspecialties, and data types. Training and testing were performed in a 3200:800 split dataset. Metrics and classes are shown in *Table 3 and 4-*

**Table 3.** evaluation metrics for named entity recognition (NER) model, and combined NER/rules model

| **Chiu & Nichols 2016 deep learning NER performance (baseline)** |  |
| --- | --- |
| Precision (CoNLL-2003) | 0.91 |
| Recall (CoNLL-2003) | 0.92 |
| F1 (CoNLL-2003) | 0.92 |
| Precision (OntoNotes 5.0) | 0.86 |
| Recall (OntoNotes 5.0) | 0.87 |
| F1 (OntoNotes 5.0) | 0.86 |
| **NER & rules layer for clinical specialty recognition** |  |
| Precision | 0.93 |
| Recall | 0.97 |
| F1 | 0.95 |
| **NER & rules layer for subspecialty or disease recognition** |  |
| Precision | 0.93 |
| Recall | 0.96 |
| F1 | 0.94 |
| **NER & rules layer for algorithm recognition** |  |
| Precision | 0.99 |
| Recall | 0.99 |
| F1 | 0.99 |
| **NER & rules layer for input feature recognition** |  |
| Precision | 0.95 |
| Recall | 0.94 |
| F1 | 0.94 |

**Table 4.** evaluation metrics for BERT-PubMed characteristics multi-classifier

| BERT-PubMed characteristics classifier |  |  |  |
| --- | --- | --- | --- |
| Classification | Precision | Recall | F1-Score |
| *Algorithm* |  |  |  |
| Neural net | 0.99 | 1.00 | 1.00 |
| Support vector machine | 1.00 | 1.00 | 1.00 |
| Regression | 0.96 | 0.96 | 0.96 |
| Decision trees | 0.97 | 0.99 | 0.98 |
| *Feature or data type* |  |  |  |
| X-ray | 1.00 | 0.99 | 1.00 |
| Computed tomography | 0.98 | 0.96 | 0.97 |
| Magnetic resonance imaging | 0.99 | 1.00 | 0.99 |
| Electroencephalogram | 0.99 | 1.00 | 1.00 |
| Electrocardiogram | 0.98 | 0.99 | 0.98 |
| Electromyogram | 0.99 | 1.00 | 0.99 |
| Ultrasound | 0.97 | 1.00 | 0.99 |
| Echocardiogram | 1.00 | 0.97 | 0.98 |
| Histology | 0.95 | 0.89 | 0.92 |
| Optical coherence tomography | 0.99 | 0.99 | 0.99 |
| Mammography | 1.00 | 1.00 | 1.00 |
| Fibreoptic endoscopy | 1.00 | 0.98 | 0.99 |
| Genomics | 0.94 | 0.99 | 0.97 |
| Biomarkers and laboratory | 0.96 | 0.95 | 0.96 |
| Natural language processing | 0.99 | 1.00 | 0.99 |
| Electronic health record data | 0.94 | 0.98 | 0.96 |
| *Clinical Specialty* |  |  |  |
| Oncology | 0.98 | 1.00 | 0.99 |
| Neurosciences | 0.99 | 0.99 | 0.99 |
| Cardiovascular | 0.97 | 0.99 | 0.98 |
| Respiratory | 0.98 | 0.99 | 0.99 |
| Gastrointestinal (luminal) | 0.96 | 1.00 | 0.98 |
| Hepatobiliary | 0.99 | 0.98 | 0.99 |
| Infectious disease | 0.97 | 0.98 | 0.98 |
| Psychiatry | 0.98 | 0.98 | 0.98 |
| Musculoskeletal | 0.90 | 0.94 | 0.92 |
| Urology | 1.00 | 0.98 | 0.99 |
| Haematology | 0.97 | 0.96 | 0.97 |
| Obstetrics and Gynaecology | 0.96 | 0.94 | 0.95 |
| Renal medicine | 0.98 | 0.99 | 0.99 |
| Intensive care | 0.97 | 0.98 | 0.98 |
| Emergency care | 0.94 | 0.97 | 0.95 |
| Paediatrics | 1.00 | 0.98 | 0.99 |
| *Clinical subspecialty / disease area* |  |  |  |
| Diabetes mellitus | 0.99 | 0.99 | 0.99 |
| Sepsis | 0.95 | 0.97 | 0.96 |
| Coronavirus disease 2019 | 0.99 | 1.00 | 1.00 |
| Skin cancer | 1.00 | 0.98 | 0.99 |
| Lung cancer | 0.96 | 0.99 | 0.98 |
| Brain cancer | 0.99 | 0.98 | 0.98 |
| Gastrointestinal cancer | 0.85 | 0.98 | 0.91 |
| Hepatobiliary cancer | 0.92 | 1.00 | 0.96 |
| Prostate cancer | 0.96 | 0.99 | 0.97 |
| Gynae-oncology | 1.00 | 0.97 | 0.99 |
| Haem-oncology | 0.96 | 0.97 | 0.96 |
| Breast cancer | 0.98 | 0.99 | 0.99 |
| Pneumonia | 0.92 | 0.98 | 0.95 |
| Epilepsy | 1.00 | 1.00 | 1.00 |
| Stroke or haemorrhage | 0.96 | 0.90 | 0.93 |
| Dementia | 0.99 | 0.99 | 0.99 |
| Ischaemic heart disease | 0.93 | 0.92 | 0.93 |
| Heart failure | 0.81 | 0.86 | 0.84 |
| Arrhythmia | 0.95 | 0.89 | 0.92 |
| Retinopathy | 0.95 | 0.98 | 0.97 |
| *Summary statistics* |  |  |  |
| Macro | 0.97 | 0.98 | 0.97 |
| Weighted | 0.98 | 0.98 | 0.98 |

### Dashboard deployment and hosting

The pipeline is deployed to the Google Cloud platform (https://cloud.google.com/) as Cloud Functions, triggered every 24-hours to discover new papers indexed in the preceding period. Due to the size of models and time required to label abstracts, the pipeline is split into two functions which share data via Google Cloud platform Pub/Sub. Google BigQuery (https://cloud.google.com/bigquery) is used to store scraped and labelled abstracts.

Deployed pipeline code hosted at: https://github.com/whizzlab/health_ai_online_pipeline

## Results

### Performance

The final pipeline (*Figure 1)* performs a sequence of tasks, feeding into the dashboard. It uses broad AI-related search terms to identify and extract abstracts and metadata from MEDLINE, before performing geolocation on affiliations. It then runs through NLP tasks that (1) label abstracts for inclusion if they represent original research that develop clinical AI models; (2) identify research at a ‘mature’ development stage which describe either evaluation of an AI algorithm vs clinicians, or prospective real-world testing; (3) labels abstracts for research characteristics including clinical specialty, subspecialty, disease, input data type, and algorithm.

In prospective evaluation of ability to correctly classify publications indexed on MEDLINE after September 2021, the inclusion classifier (task 1) achieves an Fl of 0.96 and a Matthews correlation coefficient (MCC) of 0.94. The maturity classifier (task 2) achieves an Fl of 0.91 and MCC of 0.90. The multi-class classifier (task 3) for abstract characteristics achieves a macro-average Fl of 0.97 across classes. When evaluated against a recent systematic review of deep learning^14^, out of 446 publications identified by review authors, the pipeline correctly included 438 (98.2%).

Compared to a systematic review of comparative studies^18^, the pipeline correctly labelled 81 out of 83 (97.5%) for maturity. Detailed performance metrics are reported in *Tables 1-4*

### Dashboard and global AI landscape

The interactive dashboard updates every 24 hours (https://aiforhealth.app), creating a live view of the current state of global AI. Dashboard datasets allow any cross-section of attributes to be extracted, compared, and analysed longitudinally.

To illustrate utility, we examined the entire research landscape before October 2021. Growth in clinical AI research became explainable by exponential growth functions from 2016, in total research (R2=0.999, p<0.001) and mature research (R2=0.998, p<0.001). We discovered 34178 examples of original AI research, with 1562 studies employing mature validation methods. Development and maturity heterogeneity across major themes over the past decade is illustrated in *Figure 3*. Lung, breast cancer, and retinopathy demonstrate substantial maturity relative to total research production, while cardiovascular, psychiatry, and infectious disease prediction lag behind. The distribution of data type usage across major subspecialties are shown in *Figure 4* as heatmaps, demonstrating prevalence of mature validation using radiomics (and other imaging tasks) across all AI themes. Notably, only 1.3% of all research, and 0.6% of mature research, involved an author from a low to low-middle income country (per World Bank definitions), with 93.6% of such research published after 2016 *(Figure 5)*.

**Figure 3.**
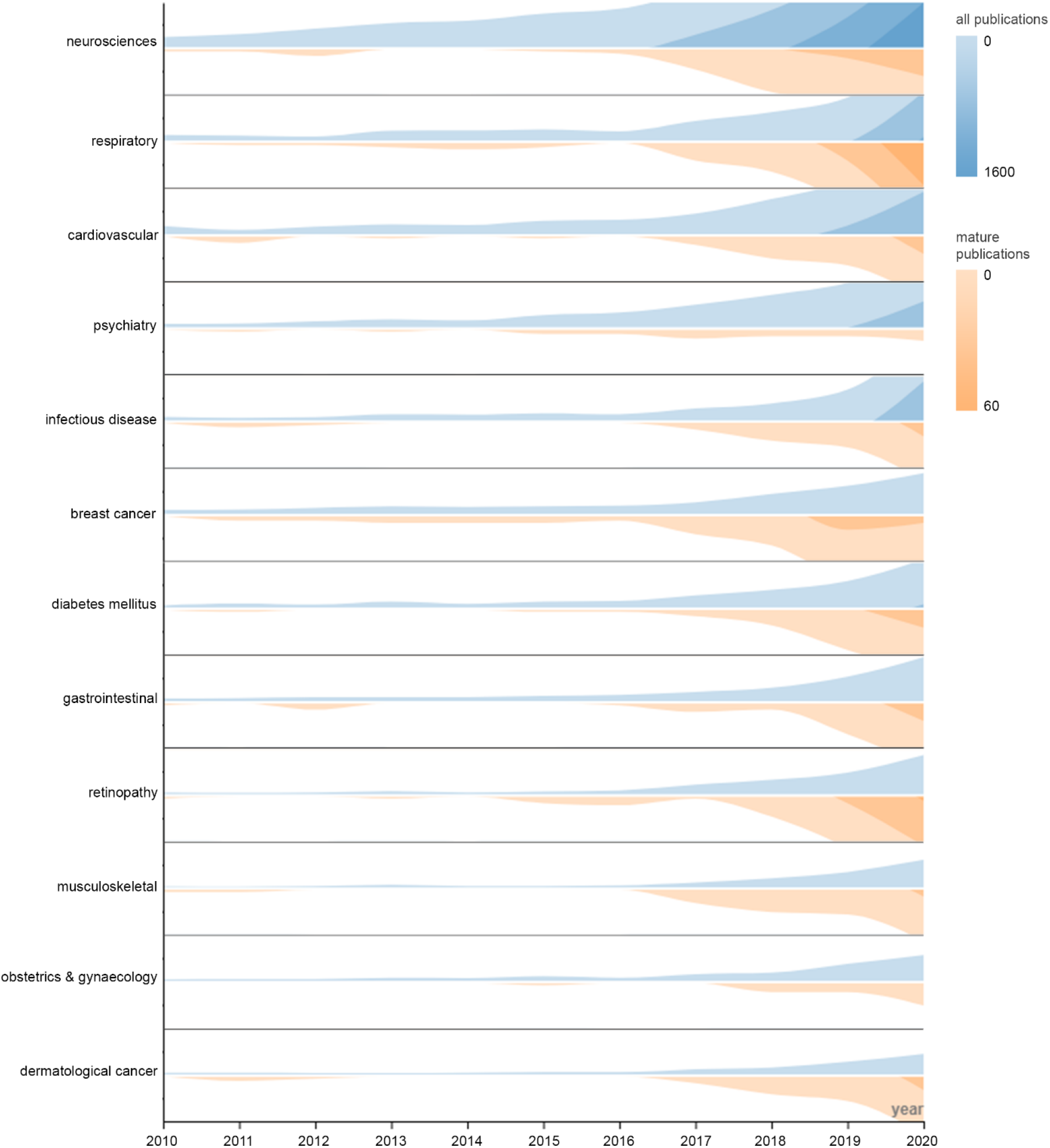
Horizon charts of relative research output across major specialties and disease areas for all publications (blue) and mature publications (orange), for each year in the past decade. Colour density is used in addition to height to represent size, maximising use of available space. Design and interpretation as described by Heer et al^21^.

**Figure 4.**
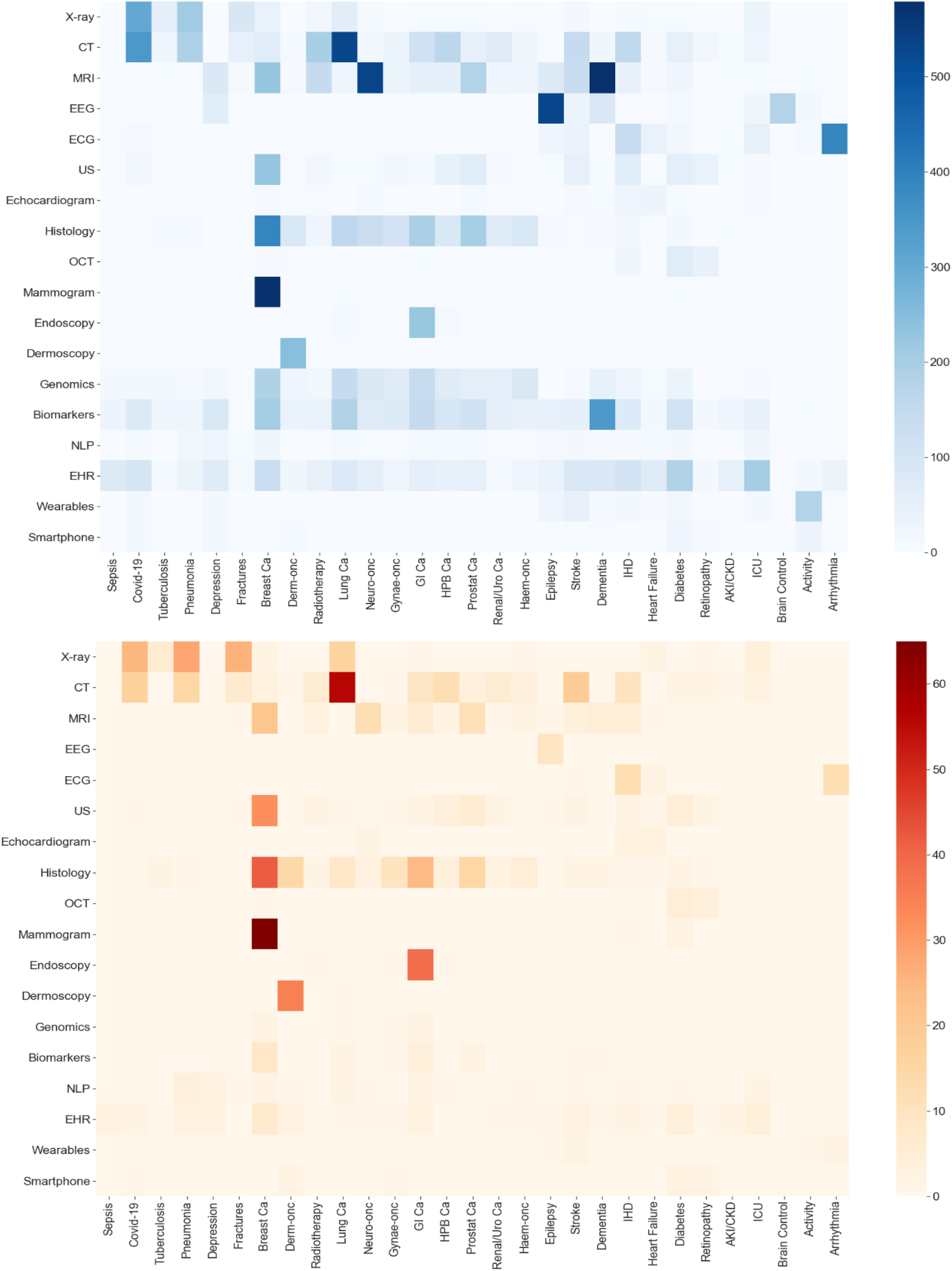
heatmap showing use of input features across subspecialty/disease areas in all (top,blue) and mature (bottom, orange) studies.

**Figure 5.**
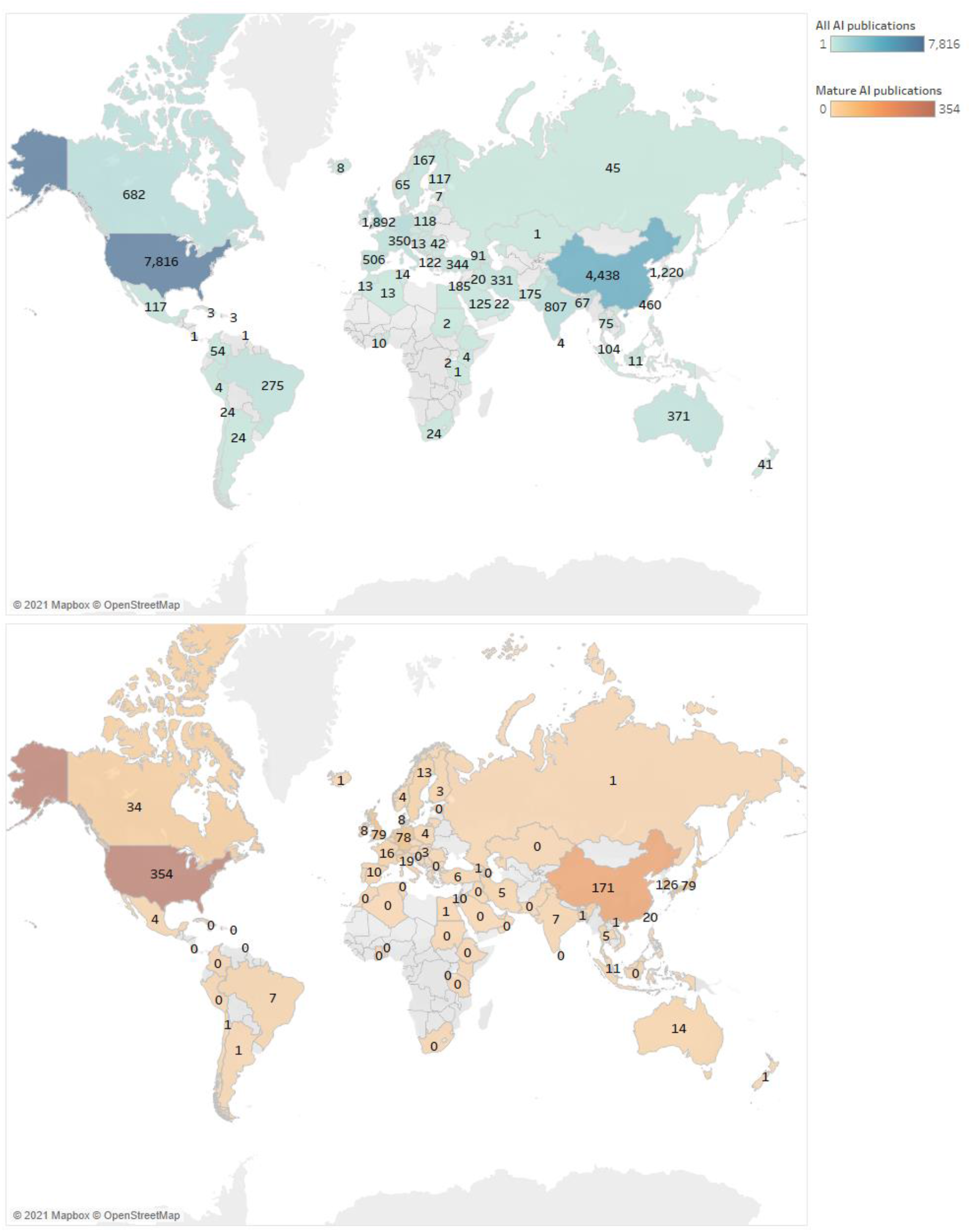
global distribution of clinical artificial intelligence research from 1998 to 2021 by first author, showing all publications (blue, top) and mature publications (orange, below). Where first author affiliation is not available, last author affiliation is used instead.

## Discussion

Previous bibliometric analyses of AI literature have relied on keywords which provide poor specificity, with significant limitations in scope of data that can be extracted from the literature^19,20^. In real-world deployment, our pipeline excels at identifying original AI research, and mature AI model development, with high specificity. The pipeline labels detailed characteristics, allowing longitudinal observation and analysis of research production and development maturity, across geography, specialties, and data types.

While demonstrating state-of-the-art NLP performance, classifier limitations include imperfect accuracy compared to careful human reviewers. This is the trade-off against time required for substantial manual characterisation. Additionally, we utilise only MEDLINE due to their supplied application programming interface (API). Finally, using text from full articles could increase classifier performance, but this was hindered by paywalled access to most publications.

We plan to continue enhancement of this resource. Code and data are made public (https://github.com/whizzlab), with the hope that function can be expanded with input from the global AI community.

## Data Availability

All data and code involved in this study are available in the GitHub repositories described within the paper.

https://github.com/whizzlab

## Acknowledgements

Thanks to the team at SparkNLP for academic licensing rights to use their named entity recognition engine.

## Data Availability

Code, data, and models are hosted online (https://github.com/whizzlab) under an open-source license.

